# Exploring predictive frameworks for malaria in Burundi

**DOI:** 10.1101/2021.07.20.21260697

**Authors:** Lionel Divin Mfisimana, Emile Nibayisabe, Kingsley Badu, David Niyukuri

**Author notes:** Corresponding author Email address (David Niyukuri). These authors contributed equally to this work (Lionel Divin Mfisimana, Emile Nibayisabe).

## Abstract

Malaria is a major public health concern in Burundi. The infection has been increasing in the last decade despite efforts to increase access to health services, and the deployment of several intervention programs. The use of different data sources can help to build predictive models of malaria cases in different sub-populations. We built predictive frameworks using generalized linear model (GLM), and artificial neural network to predict malaria cases in four sub-populations (pregnant women and children under 5 years, pregnant women, children between 0 and 11 months, children between 12 and 59 months), and the overall general population. The results showed that almost half malaria infections are observed in pregnant women and children under 5 years, but children between 12 and 59 months carry the highest burden. Neural network model performed better in predicting total cases compared to GLM. But the latter provided information on the effect of predictors, which is an important source of information to mainstream target interventions. Early prediction of cases can provide timely information needed to be proactive for intervention strategies, and it can help to mitigate the epidemics and reduce its impact on populations and the economy.

## 1. Introduction

Malaria infection has been, and still a major public health concern in Burundi [1]. The World Health Organization (WHO) estimates show that more than 80% of the Burundian population are at high risk of acquiring malaria infection [1, 2]. Malaria cases increased from 2.6 million in 2013 to 8.3 million in 2016 with a further 18 percent increase in the first half of 2017 compared to 2016 [3]. In 2017 malaria was declared a national epidemic [4, 5, 6, 7]. According to the Institute for Health Metrics and Evaluation (IHME), in 2019 malaria was ranked fourth cause of death after tuberculosis in Burundi [8, 9].

The health system still has many challenges, despite efforts by the Government to improve by increasing access to healthcare facilities and technical capacities [10]. Burundi suffers from a shortage of qualified personnel, medical resources, and high burden of diseases [11]. According to the World Bank statistics in 2017, Burundi was estimated to have 0.1001 physicians for 1000 people [12], and in 2010, the density of doctors, nurses and midwives combined was estimated to be 2 per 10,000 population [13].

In a study published in 2011 [14], the authors claimed that 17.4% of patients did not have access to health care at all, while 81.5% of patients were forced to go into debt or sell their property to pay for the health costs. Furthermore, the authors of same study [14], and the report of health systems [11] reported a huge disparity between the urban areas especially the economic capital Bujumbura, and the other parts of the country.

According to the World Vision International, an international Non Government Organization (NGO) which support malaria interventions in Burundi, some major drivers of malaria infection in Burundi are: the low usage of Insecticide Treated Bed Nets (ITNs), climate change, population density, shifting of farming practices, food insecurity, and lack of knowledge and action to prevent malaria in the communities [15].

Studies have shown association of malaria cases with temperature, humidity, rainfall, usage of mosquito nets, geographical location, and socioeconomic factors [16, 17, 18, 19]. This means that the data of the above mentioned factors can be used to predict number of malaria cases at a certain level of certainty. A study in Uganda showed that malaria is highly endemic with temperatures and precipitation that allow stable transmission throughout the year with relatively low seasonal variability in most parts of the country [20, 21]. In areas with a long dry season such as the Sahel, and particularly in Burkina Faso, rains determine the abundance of mosquito populations and thus the intensity of transmission with mosquitoes then benefiting from multiple puddles to reproduce and also high humidity levels of the surrounding air. In Niger, a 16% increase in rainfall between 2005 and 2006 was accompanied by a 132% increase in mosquito abundance in the village of Banizoumbou, located in the Sahelian zone [22]. The impact of altitude on malaria transmission is directly related to the decrease in temperature that influences both the vector (anopheles mosquito) and the parasite [23, 24]. But also the wind speed varies according to the altitude so that at high altitude mosquitoes may lose their vectorial capacity.

Using multiple data sources for malaria prediction has been argued to complement the surveillance system of malaria [25] which is often based on reported data. Therefore, given the current knowledge on factors which may increase or decrease malaria prevalence, it is important to enhance decision making, and resource allocation with targeted interventions for effective malaria control. And since there is no one-size fits all intervention to eliminate malaria, modelling frameworks can help to explore a range of interventions given different predictions of infection cases, epidemic estimates, and cost effectiveness. This is crucial for decision making to be able to tail interventions and optimize resources as stated in the the WHO’s Global Technical Strategy for Malaria 2016–2030 [26].

The objective of this study was to use available data sources which have been proven to be associated to malaria infection [27, 28, 29, 19] to build basic predictive frameworks to predict malaria cases in different sub-populations in Burundi, namely response variables: pregnant women and children under 5 years, among pregnant women, children between 0 and 11 months, children between 12 and 59 months, and the overall general population. This provides the starting point for more complex models with granular data to understand the transmission dynamics of malaria, and subsequent control measures in Burundi.

## 2. Methods

### 2.1. Data

The study was conducted using monthly data collected in from different provinces of Burundi for the period 2010–2017. The data consists of records of total malaria cases, cases among children under 5 years and pregnant women, as well as mosquito nets distributed to children and pregnant women. We also used monthly weather data. We used also annual population estimates and schooling rates for all provinces.

A record of the total nationwide malaria cases was available from the National Integrated Malaria Control Program of the Ministry of Health. Detailed monthly data sets on malaria infection for children (from 0 to 11 months then up 12 to 59 months male and female), and pregnant women (before and after first trimester of pregnancy), the number of mosquito nets distributed (for children under five years and pregnant women) per each province were available from the Department of National System of Health Information of the Ministry of Health.

Geographical, and monthly climate data (weather) per province were provided by the Geographical Institute of Burundi. In addition, demographic data for each province were provided by the open library of the National Institute of Statistics. And the schooling data per province per year were provided by the Planning Office of the Ministry of Education and Scientific Research.

### 2.2. Data imputation

We combined all the databases in one data frame, after cleaning, we had a new data frame with the following variables: area, longitude, latitude, altitude, rainfall, temperature, humidity, population, education, malaria cases among children under 11 months, malaria cases among children between 12 and 59 months, malaria cases among pregnant women with less than 3 months, malaria cases among pregnant women beyond 3 months, bed-nets given to children, bed-nets given to pregnant women, severe malaria cases, and other malaria cases.

Climate data had missing values, and to deal with these we used a nonparametric missing value imputation approach using random forest method (missForest) [30]. This method is one the methods often used to impute continuous and/or categorical data including complex interactions and nonlinear relations.

Based on correlation analysis results, to build the predictive models for malaria cases, we used only some variables: altitude, rainfall, temperature, humidity, population density, education level, number of pregnant women, number of children under five years, and number of bed-nets distributed. We built the models using randomly 75% of the data set for training, and 25% for testing. But in order to be more realistic, we also considered a scenario where we used data data from 2010 up to 2016 for training, and predict 2017 cases. For that scenario we considered only predicting the monthly total cases of malaria.

### 2.3. Data exploration

With the monthly data for a period of eight years (2010 - 2017), we explored distributions of malaria cases within different sub-populations in different provinces. We looked at the differences of trends among the 8 years by using Wilcoxon Rank Sum Test [31] to investigate if there has been distribution (median) difference between monthly cases among pregnant women and children under 5 years along the years. The use of the Wilcoxon test tells us whether the median values of the two-by-two comparison of the monthly cases were from same continuous distribution or not. The null hypothesis is that the vectors of monthly cases were from the same distribution. This was rejected when the p-value was less than the 0.05 significance level.

### 2.4. Modelling: General Linear Model and Neural Network

We built a general linear model (GLM) [32], and an artificial neural network model (ANN) [33] to predict malaria cases in different scenarios. Data were normalized for the neural network model. Each response variable (malaria cases among children under 5 years, malaria cases among pregnant women, malaria cases among children between 0 and 11 months, malaria cases among children 12 and 59 months, and overall total malaria cases) was evaluated by both types of models. To ensure model validity and performance, we used cross-validation approach with 10 fold replication. And to assess the goodness of fit, we computed the mean square root error (MSRE) of the difference between true and predicted values of malaria cases.

The generalized linear model (GLM) is a flexible generalization of ordinary linear regression that allow the dependent variable to be non-normal. Given *Y* dependant variables, and *X* independent variables. The expected value of *Y* is given by

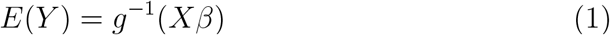

where *g* is called the link function, and *β* are linear combination parameters. In our GLM model, the link function was a Gaussian distribution. The GLM generalizes linear regression by allowing the linear model to be related to the response variable via a link function and by allowing the magnitude of the variance of each measurement to be a function of its predicted value. In our model, the link function was the function gamma. With GLM fitting to data, we get effect of each predictor to the response (output) with assumption that other predictors are omitted. To be able to see the effects of the all predictors we performed an ANOVA to the GLM outputs with Chi-Square test which consider adding sequentially terms (predictors) to the model.

Artificial neural networks are tools used in machine learning which are able to detect multiple nonlinear interactions among a series of input variables (predictors). They are brain-inspired systems which are intended to replicate the way that we humans learn. Neural networks consist of input and output layers, as well as a hidden layer consisting of units that transform the inputs into a sable form for the output layer. After exploration of different artificial neural network configurations with number of layers and neurons, we considered an artificial neural network with two hidden layers with respective internal nodes (3, 4), and the neural network was trained 10 times using *neuralnet* package [34].

For any of the above mentioned modeling approaches, the goodness of fit was measured by the Root Mean Square Error (RMSE):

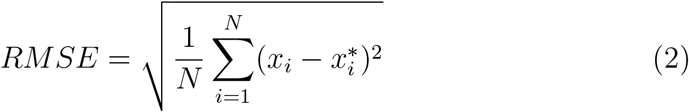

where *x*_*i*_ is the observed number of cases and 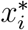 the predicted number of cases, and the coefficient of variation (CV). To evaluate the models, we used cross-validation, we re-sampled 10 times the data and run the model, and we summarized the model performance by RMSE. All the computations were performed using R (version 3.4.4) software [35] in a Linux environment.

In our models, the outcome of interest was the number of malaria cases: the overall total cases, cases among pregnant women and children under 5 years, cases among pregnant women, cases among children between 0 and 11 months, and those between 12 and 59 months.

## 3. Results

### 3.1. Descriptive analysis

A descriptive analysis of annual malaria cases shows that the epidemic curves for the five categories between 2010 and 2017 have an increasing trend as we can see at Figure 1. The epidemic curves show what is well known that pregnant women and children under 5 years are at risk for malaria infection, they are the first group with high number of cases. However, although we have a lot of malaria cases for pregnant women and children under 5 years, almost a half of cases came from other populations groups of adults and children above 5 years.

**Figure 1:**
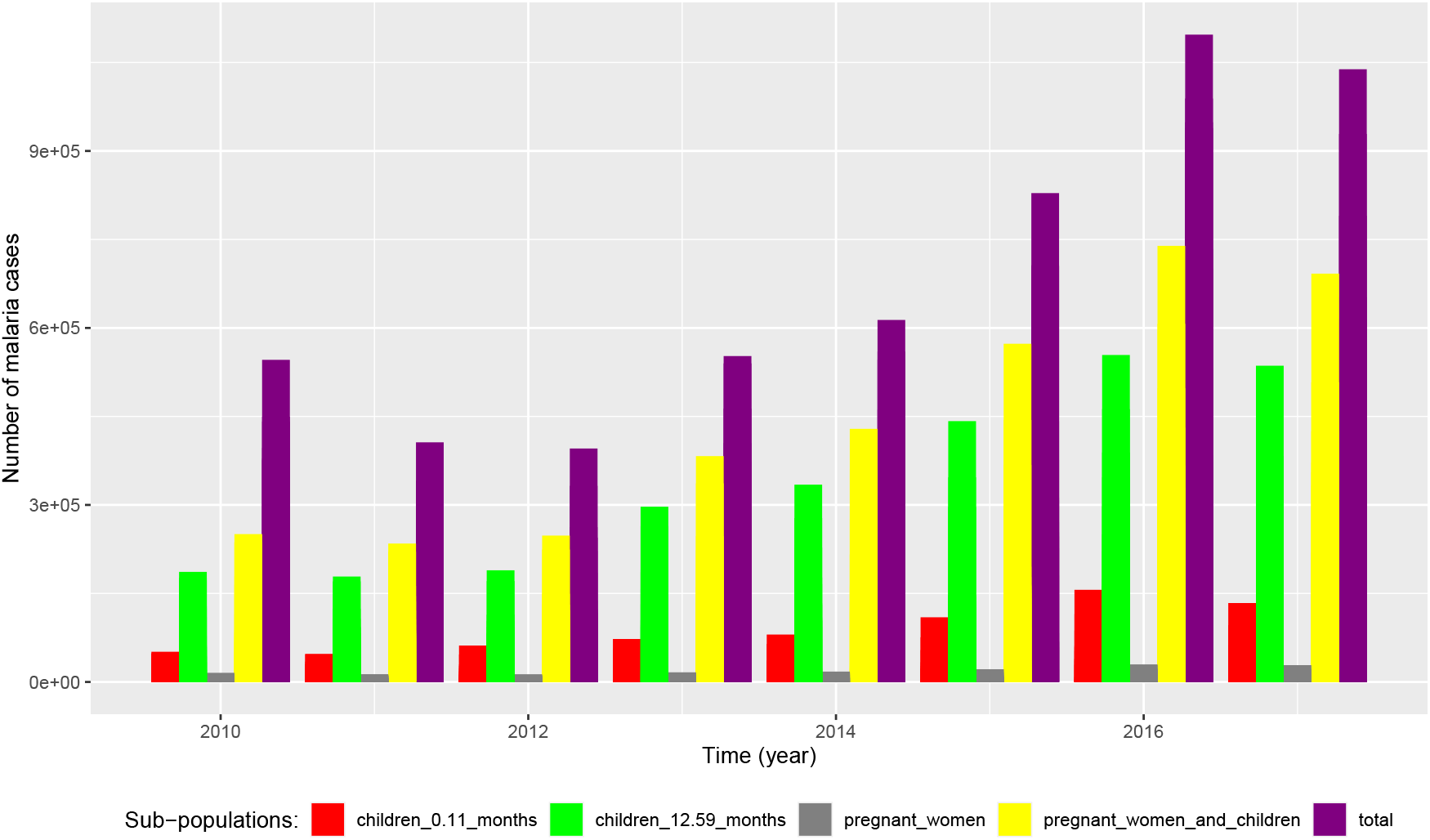
Country level malaria cases in children between 0 and 11 months, children between 12 and 59 months, pregnant women, pregnant women and children under 5 years, and the overall total cases between 2010 and 2017.

But when this high risk group is split in three subgroups with pregnant women, children between 0 and 11 months, children between 12 and 59 months, we can see that children between 12 and 59 months have a disproportionate burden of malaria infection compared to other subgroups in all the eight years (2010 - 2017), with pregnant women group being the one with low cases. Regarding geographical distribution, Figure 2 shows that provinces of Kirundo, Gitega, Muyinga, and Ngozi were much affected by the infection compared to others.

**Figure 2:**
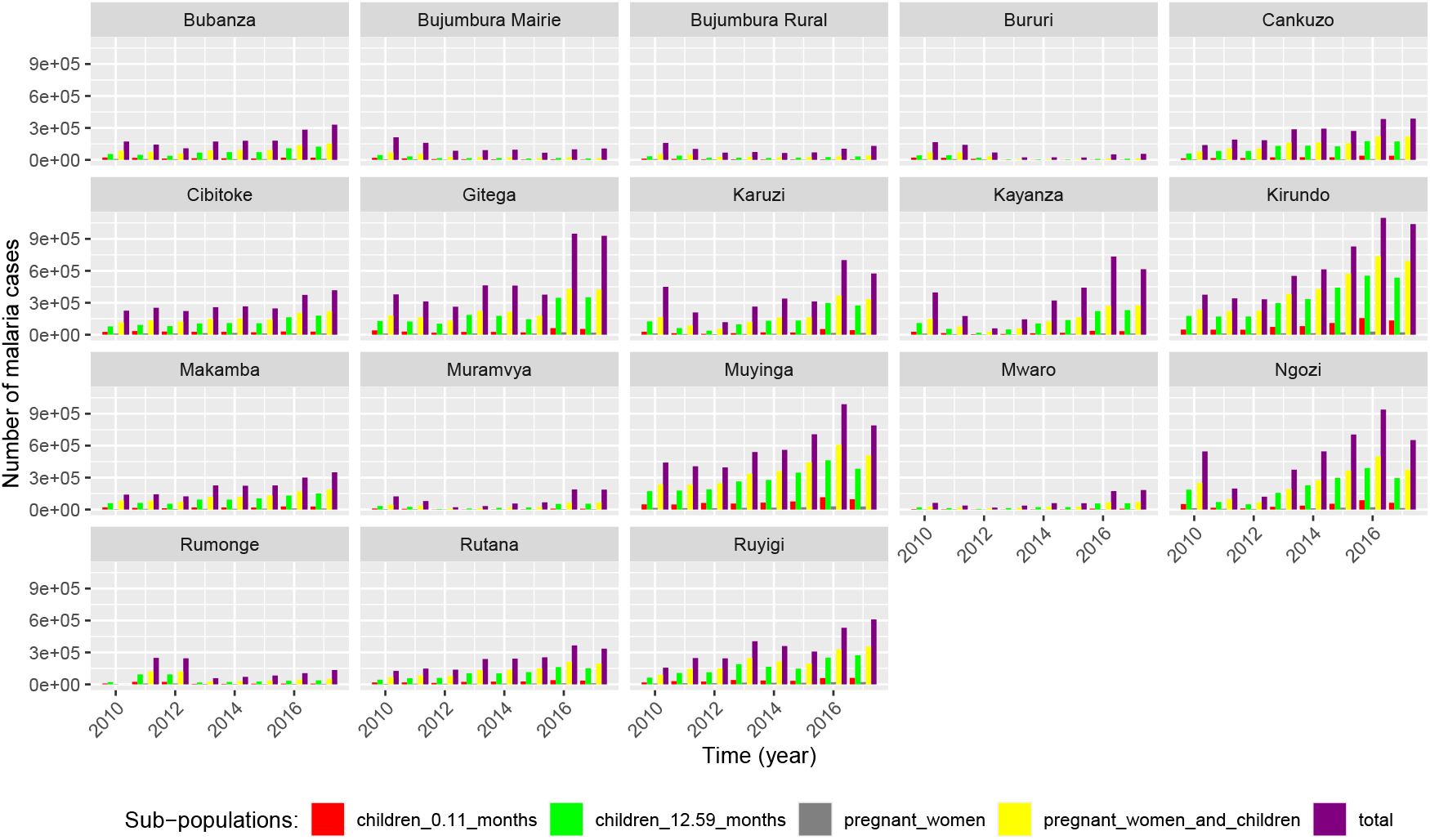
Province level malaria cases in children between 0 and 11 months, children between 12 and 59 months, pregnant women, pregnant women and children under 5 years, and the overall total cases between 2010 and 2017.

When we aggregate monthly the number of malaria cases for the eight years (2010 - 2017), the Figure 3 shows that the total malaria cases have same trend as cases among pregnant women and children under 5 years, and cases among children between 12 and 59 months. We can say that the trend of pregnant women and children under 5 years can benchmark malaria epidemic. However, children between 12 and 59 months have same trend as cases pregnant women and children under 5 years, but this is explained by the fact the large proportion of the late category is made of children between 12 and 59 months.

**Figure 3:**
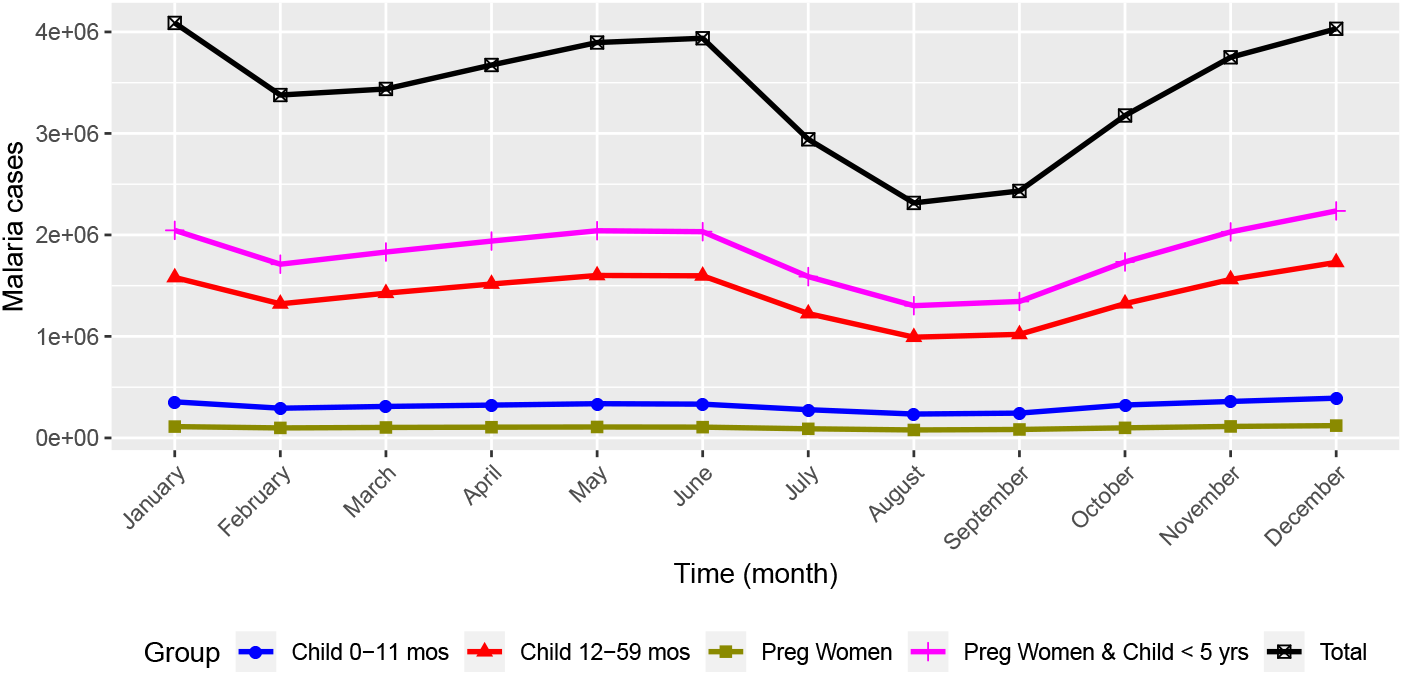
Aggregating monthly malaria cases in different sub-populations between 2010 and 2017.

### 3.2. Outbreaks comparison

As we said in subsection 3.1, in the 8 years period (2010 – 2017), the epidemic curve of malaria cases has an increasing trend. Considering malaria cases in pregnant women and children under 5 years as a benchmark of the trend, if we compare the monthly number of cases in those 8 years, Figure 4 shows that although the magnitude is different, the overall trend across months is similar of what we see at Figure 3. In January, we have high number of infections which decreases in February and start increasing by March up to June where an sharp decrease is observed in summer with lowest cases in August. In September, cases start increasing again up to December.

**Figure 4:**
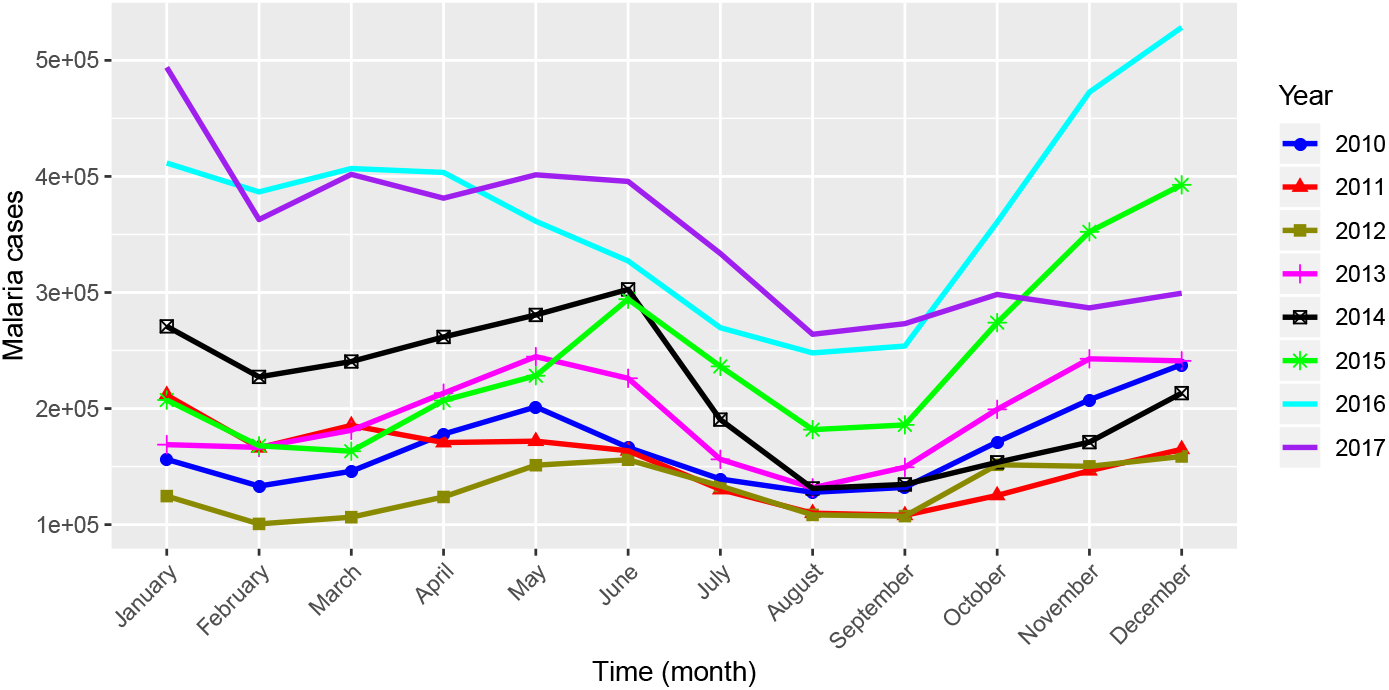
Monthly malaria cases in pregnant women and children under 5 years in 8 years periods (2010 - 2017)

Across the years, monthly data were different, even the year with the highest number of cases (2016), it did not have high cases in all months. A comparison between monthly cases among pregnant women and children under 5 years in the 8 years (2010 - 2017) using Wilcoxon Rank Sum Test showed that there has been much disparity since in the Table 1, thus, the median values of malaria cases among pregnant women and children under 5 years were from different distribution in many cases, with a p-value less than 5%. But we have few scenarios where we could find that the median values of monthly cases were from same distribution. That could be seen in the following years: between 2010 and 2011, 2011 and 2012, 2013 and 2014, and 2016 and 2017.

**Table 1:** P-values for the Wilcoxon Rank Sum Test between monthly number of malaria cases among pregnant women and children under 5 years.

| Year | 2010 | 2011 | 2012 | 2013 | 2014 | 2015 | 2016 | 2017 |
| --- | --- | --- | --- | --- | --- | --- | --- | --- |
| 2010 | NA | 0.3010 | < 0.001 | < 0.001 | 0.0340 | < 0.001 | < 0.001 | < 0.001 |
| 2011 | 0.3010 | NA | 0.0640 | 0.0160 | < 0.001 | 0.0050 | < 0.001 | < 0.001 |
| 2012 | < 0.001 | 0.0640 | NA | < 0.001 | < 0.001 | < 0.001 | < 0.001 | < 0.001 |
| 2013 | < 0.001 | 0.0160 | < 0.001 | NA | 0.204 | 0.0160 | < 0.001 | < 0.001 |
| 2014 | 0.0340 | < 0.001 | < 0.001 | 0.204 | NA | 0.8500 | < 0.001 | < 0.001 |
| 2015 | < 0.001 | 0.0050 | < 0.001 | 0.0160 | 0.85 | NA | < 0.001 | 0.0090 |
| 2016 | < 0.001 | < 0.001 | < 0.001 | < 0.001 | < 0.001 | < 0.001 | NA | 0.9700 |
| 2017 | < 0.001 | < 0.001 | < 0.001 | < 0.001 | < 0.001 | 0.009 | 0.9700 | NA |

### 3.3. Models outputs

The following results in Table 2 are the error values (MSRE) for the GLM and ANN models outputs compared to observed data for malaria cases in the overall population (total cases), among pregnant women and children under 5 years, among pregnant women, among children between 0 and 11 months, and those between 12 and 59 months. ANN performs better in estimating total malaria cases compared to GLM model (Table 2). But the generalized linear models do not perform too bad since the difference between generalized linear model and neural network model is between two and three thousands cases (CV and MSRE). But for cases among pregnant women, and children,the generalised linear model performed better compared to neural network model (Table 2).

**Table 2:** Mean Square Root Error and Cross Validation between observed and predicted values of malaria cases with generalized linear model and artifical neural network.

| Malaria cases | MSRE (GLM) | CV (GLM) | MSRE (ANN) | CV (ANN) |
| --- | --- | --- | --- | --- |
| Total | 7810 | 6019 | 4125 | 4024 |
| Pregnant women & children (< 5 years) | 141 | 227 | 460 | 488 |
| Pregnant women | 26 | 22 | 25 | 28 |
| Children (0 and 11 months) | 592 | 661 | 511 | 560 |
| Children (12 and 59 months) | 536 | 721 | 655 | 737 |

Focusing on GLM model, Table 3 shows that compared to others response variables, the overall total number of malaria cases was the only response variable where all parameters estimates associated to predictors have statistically significant (based on their p-values) effect on the outcome.

**Table 3:**
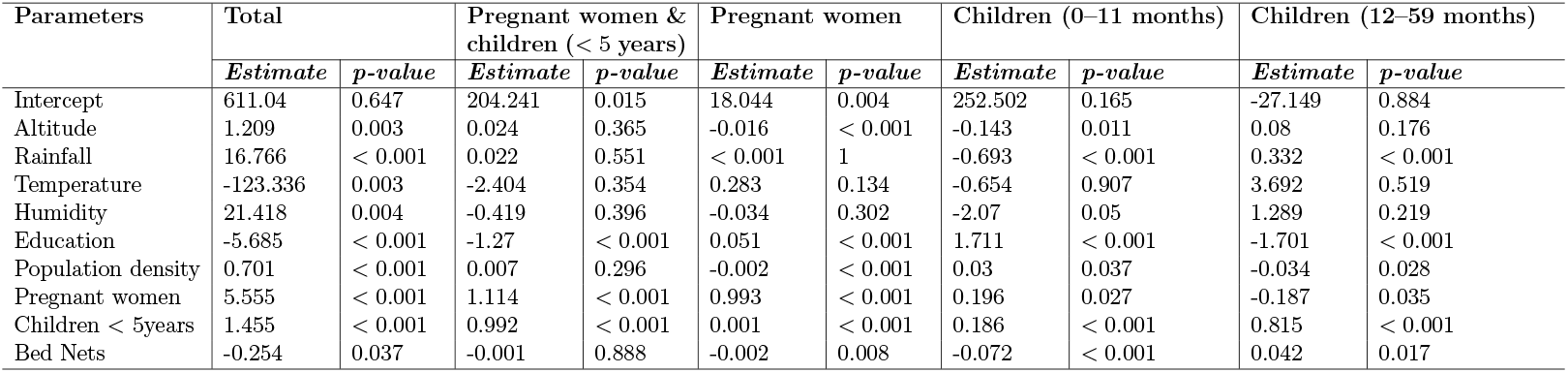
Parameter estimates and p-values of the generalised linear model for five subpopulations of malaria cases

| Parameters | Total |  | Pregnant women & children (< 5 years) |  | Pregnant women |  | Children (0–11 months) |  | Children (12–59 months) |  |
| --- | --- | --- | --- | --- | --- | --- | --- | --- | --- | --- |
|  | *Estimate* | *p-value* | *Estimate* | *p-value* | *Estimate* | *p-value* | *Estimate* | *p-value* | *Estimate* | *p-value* |
| Intercept | 611.04 | 0.647 | 204.241 | 0.015 | 18.044 | 0.004 | 252.502 | 0.165 | -27.149 | 0.884 |
| Altitude | 1.209 | 0.003 | 0.024 | 0.365 | -0.016 | < 0.001 | -0.143 | 0.011 | 0.08 | 0.176 |
| Rainfall | 16.766 | < 0.001 | 0.022 | 0.551 | < 0.001 | 1 | -0.693 | < 0.001 | 0.332 | < 0.001 |
| Temperature | -123.336 | 0.003 | -2.404 | 0.354 | 0.283 | 0.134 | -0.654 | 0.907 | 3.692 | 0.519 |
| Humidity | 21.418 | 0.004 | -0.419 | 0.396 | -0.034 | 0.302 | -2.07 | 0.05 | 1.289 | 0.219 |
| Education | -5.685 | < 0.001 | -1.27 | < 0.001 | 0.051 | < 0.001 | 1.711 | < 0.001 | -1.701 | < 0.001 |
| Population density | 0.701 | < 0.001 | 0.007 | 0.296 | -0.002 | < 0.001 | 0.03 | 0.037 | -0.034 | 0.028 |
| Pregnant women | 5.555 | < 0.001 | 1.114 | < 0.001 | 0.993 | < 0.001 | 0.196 | 0.027 | -0.187 | 0.035 |
| Children < 5years | 1.455 | < 0.001 | 0.992 | < 0.001 | 0.001 | < 0.001 | 0.186 | < 0.001 | 0.815 | < 0.001 |
| Bed Nets | -0.254 | 0.037 | -0.001 | 0.888 | -0.002 | 0.008 | -0.072 | < 0.001 | 0.042 | 0.017 |

Thus, by sequentially adding one predictive variable for all the generalised models in the table 3, all the predictors had significantly effect to the outcomes. Some of the variables have an increase or decrease effect on malaria cases. Rainfall, humidity, number of pregnant women, and the number of children under five years have an increase effect on the overall malaria cases. But, education, temperature, and the number of bed nets distributed to pregnant women and children have a decreasing effect on the overall malaria cases.

The total monthly predicted malaria cases for 2017 (Figure 5) was 7.73 millions with neural network model, and 7.47 millions with generalised linear model, but the observed data was 7.82 millions. Overall, the neural network model performed better compared to generalised linear model. But looking on months data, we can see that in some months one model would have better predictions compared to another.

**Figure 5:**
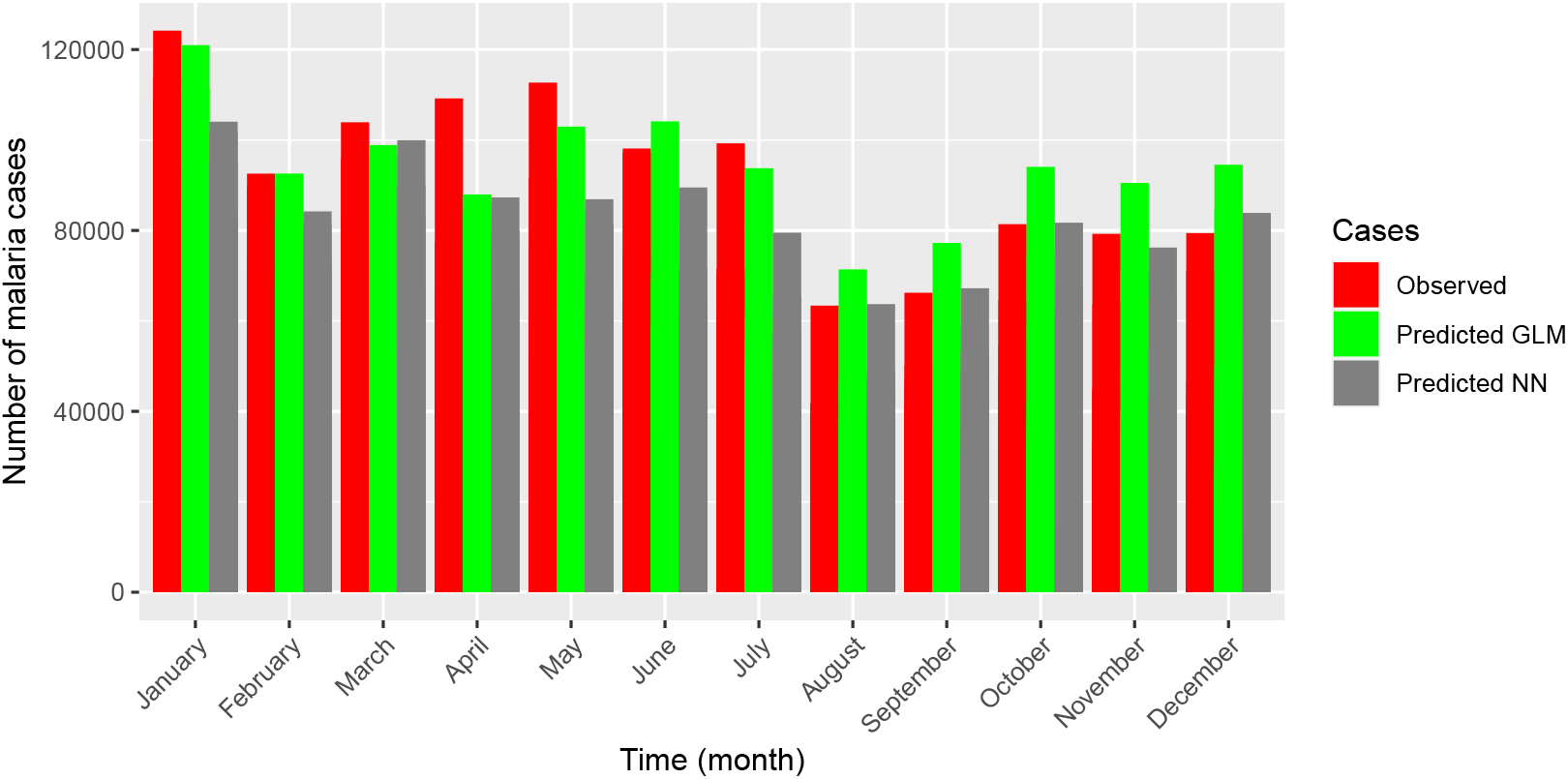
Observed and predicted monthly total malaria cases in 2017

## 4. Discussion

Splitting the population in sub-groups shed lights on more high risk groups to which interventions should be intensified. We have seen much proportion of cases were among children between 12 and 59 months. It is much likely that we may discover different transmission patterns across different age groups within other no reported groups of adults and children above 5 years. Thus, having individual level data can contribute to enhance interventions. The high burden of children between 12 and 59 months is supported by the literature which shows that new born are relatively protected for malaria infection since they can acquire immunity to malaria from their mothers who were exposed to the infection, but this immunity decrease by 6-9 months [36, 37].

It is important to note that despite higher number of malaria cases for pregnant women and children under 5 years, other sub-groups of adults and children above 5 years are also bearing non-negligible burden of malaria infection. These other groups which are suppose to have much immunity compared to pregnant women and children below 5 years play a key role in the transmission cycle of malaria. Thus, specific interventions should target these groups, and epidemiological investigations and behaviour should be conducted because this is not related to their weak immunity as pregnant women and children under five years.

Some months of the year are characterised by higher cases compared to others, and others have very low cases. This may be explained by weather changes and rains, but we have seen that the monthly trends in the 8 years were not the same, in some months there was higher cases and lower in others regardless of the overall annual cases (see Figure Figure 4). This observation is supported by the study by Nkurunziza et al in which the authors found that malaria incidence was positively associated to minimum temperature between two months, and that rainfall and maximum temperature in a given month have a possible negative effect on malaria incidence of the same month [14]. These different trends in different months show the existence of different seasons of malaria transmission in Burundi. But even if there are some months with high infection rates, same month of the years can have more or less malaria cases if we look at Figure 4. It shows that, it is not only weather changes that affect malaria cases. Other factors to explain such behaviour can be the climate change, and socio-economic factors without excluding intensity of prevention interventions such as distribution of mosquito nets, and medication. However, one of our limitation in this study, we could not get granular data at province level to understand truly the effect of socioeconomic factors and prevention interventions on malaria cases.

Generalized linear model, and the neural network both estimate number of malaria cases at a certain accuracy. The main difference between these two approaches is that GLM is more transparent than neural network. The late works like a *black-box* which detects multiple nonlinear interactions among a series of input variables before giving the output, whereas the GLM provides details on how the predictors affect the final output (response variables) through their coefficients. The neural network model works better for predicting the overall total cases of malaria. However, it does not provide information on the magnitude of effect of predictors (variables) to the outcome (malaria of cases). Thus, for planning purposes, it may be advisable to use neural network to predict overall malaria cases. But generalised linear model gives the effect of each of predictors (variables) and if it increase or decrease the likelihood of the outcome (malaria cases), thus, this helps to know where to focus more attention for interventions.

From GLM, we have seen that associated parameters to education and bed nets had negative estimates, which means they have a decrease effect on total malaria cases. Therefore, sensitization to increase awareness, and capacity building for best practices to avoid malaria infection in communities combined to messages targeting school learners and students can also help to reduce malaria infection.

Malaria still a public health challenge in Burundi, deep dive studies can help to ascertain the principal drivers of the transmission which will then present opportunities for tailor made interventions for effective malaria control. Thus, the use of models that can help to predict malaria epidemics and being proactive for interventions designs. And stratification of populations can help to identify more risky groups, and to tailor interventions. Furthermore, including many variables in the predictive frameworks can increase accuracy and enhance understanding of malaria dynamic and hence tailor adequate intervention measures.

## Data Availability

All required data are available in the manuscript

## Acknowledgments

Authors acknowledge the contribution of Emanuel Muema Dominic during data analysis and modelling, and Edmund Yamba for his valuable comments to the early version of the manuscript.

## Authors contributions

All authors conceptualized the study. EN, and LM collected and collated the data, DN wrote the code to clean, and analyze the data, DN conducted the analysis and modelling part, KB revised extensively the first draft of the manuscript. All authors wrote and reviewed the manuscript.

## References

[1] World malaria report 2016, World Health Organization (WHO), Geneva, 2017.

[2] World malaria report 2017, World Health Organization (WHO), Geneva, 2018.

[3] President’s Malaria Initiative, Malaria Operational Plan FY 2017, United States Agency for International Development (USAID), 2016.

[4] Weekly Bulletin on Outbreaks and other Emergencies: Week 27: 01– 07 July 2017, World Health Organization (WHO)/Regional Office for Africa, 2017.

[5] UNICEF Burundi Humanitarian Situation Report – 31 March 2017, United Nations For Children (UNICEF)/Burundi, 2017.

[6] Weekly Bulletin on Outbreaks and other Emergencies: Week 32: 05 – 11 August 2017, World Health Organization (WHO)/Regional Office for Africa, 2017.

[7] P. Lok, S. Dijk, Malaria outbreak in burundi reaches epidemic levels with 5.7 million infected this year, British Medical Journal Publishing Group (2019).

[8] Health metrics for Burundi, http://www.healthdata.org/burundi, 2020. Accessed: 2021-05-26.

[9] T. Vos, S. S. Lim, C. Abbafati, K. M. Abbas, M. Abbasi, M. Abbasifard, M. Abbasi-Kangevari, H. Abbastabar, F. Abd-Allah, A. Abdelalim, et al., Global burden of 369 diseases and injuries in 204 countries and territories, 1990–2019: a systematic analysis for the global burden of disease study 2019, The Lancet 396 (2020) 1204–1222.

[10] D. Sinzinkayo, D. Baza, V. Gnanguenon, C. Koepfli, The lead-up to epidemic transmission: malaria trends and control interventions in burundi 2000 to 2019, Malaria Journal 20 (2021) 1–7.

[11] R. Lozano, F. Garrido, Improving health system efficiency (health systems governance and financing), World Health Organization. file:///Users/hectorcarrasco/Downloads/WHO HIS HGF CaseStudy 15. 7 eng 20 (2015).

[12] World health organization’s global health workforce statistics, https://data.worldbank.org/indicator/SH.MED.PHYS.ZS, 2017. Accessed: 2021-05-26.

[13] Density of doctors, nurses and midwives in the 49 priority countries, https://www.who.int/hrh/fig_density.pdf?ua=1, 2010. Accessed: 2021-05-26.

[14] H. Nkurunziza, A. Gebhardt, J. Pilz, Geo-additive modelling of malaria in burundi, Malaria Journal 10 (2011) 234.

[15] Eight facts about burundi’s malaria epidemic, https://www.wvi.org/article/8-facts-about-burundis-malaria-epidemic, 2017. Accessed: 2021-05-26.

[16] T. Li, Z. Yang, M. Wang, Temperature, relative humidity and sunshine may be the effective predictors for occurrence of malaria in guangzhou, southern china, 2006–2012, Parasites & vectors 6 (2013) 155.

[17] L. M. Beck-Johnson, W. A. Nelson, K. P. Paaijmans, A. F. Read, M. B. Thomas, O. N. Bjørnstad, The effect of temperature on anopheles mosquito population dynamics and the potential for malaria transmission, PLOS one 8 (2013) e79276.

[18] M. Pascual, J. A. Ahumada, L. F. Chaves, X. Rodo, M. Bouma, Malaria resurgence in the east african highlands: temperature trends revisited, Proceedings of the National Academy of Sciences 103 (2006) 5829–5834.

[19] H. M. Semakula, G. Song, S. Zhang, S. P. Achuu, Potential of household environmental resources and practices in eliminating residual malaria transmission: a case study of tanzania, burundi, malawi and liberia, African health sciences 15 (2015) 819–827.

[20] Uganda Malaria Control Strategic Plan 2005/06 – 2009/10, Uganda Ministry of Health, Uganda, 2005.

[21] Uganda Malaria Control Strategic Plan 2014-2020, Uganda Ministry of Health, Uganda, 2014.

[22] A. Bomblies, J.-B. Duchemin, E. A. Eltahir, Hydrology of malaria: Model development and application to a sahelian village, Water Resources Research 44 (2008).

[23] M. N. Bayoh, S. W. Lindsay, Effect of temperature on the development of the aquatic stages of anopheles gambiae sensu stricto (diptera: Culicidae), Bulletin of entomological research 93 (2003) 375–381.

[24] E. K. Kipruto, A. O. Ochieng, D. N. Anyona, M. Mbalanya, E. N. Mutua, D. Onguru, I. K. Nyamongo, B. B. Estambale, Effect of climatic variability on malaria trends in baringo county, kenya, Malaria journal 16 (2017) 1–11.

[25] X. Wang, B. Yang, J. Huang, H. Chen, X. Gu, Y. Bai, Z. Du, Iasm: A system for the intelligent active surveillance of malaria, Computational and mathematical methods in medicine 2016 (2016).

[26] Global technical strategy for malaria 2016–2030, https://www.who.int/malaria/publications/atoz/9789241564991/en/, 2021. Accessed: 2021-05-26.

[27] U. Haque, T. Sunahara, M. Hashizume, T. Shields, T. Yamamoto, R. Haque, G. E. Glass, Malaria prevalence, risk factors and spatial distribution in a hilly forest area of bangladesh, PLoS One 6 (2011) e18908.

[28] N. D. Thang, A. Erhart, N. Speybroeck, L. X. Hung, C. T. Hung, P. Van Ky, M. Coosemans, U. D’Alessandro, et al., Malaria in central vietnam: analysis of risk factors by multivariate analysis and classification tree models, Malaria Journal 7 (2008) 28.

[29] E. A. Onyango, O. Sahin, A. Awiti, C. Chu, B. Mackey, An integrated risk and vulnerability assessment framework for climate change and malaria transmission in east africa, Malaria journal 15 (2016) 551.

[30] D. J. Stekhoven, missforest: Nonparametric missing value imputation using random forest, Astrophysics Source Code Library (2015).

[31] A. Imam, U. Mohammed, C. Moses Abanyam, On consistency and limitation of paired t-test, sign and wilcoxon sign rank test, IOSR Journal of Mathematics 10 (2014) 1–6.

[32] D. Bates, M. Maechler, B. Bolker, S. Walker, et al., lme4: Linear mixed-effects models using eigen and s4, R package version 1 (2014) 1–23.

[33] K. Gurney, An introduction to neural networks, CRC press, 1997.

[34] F. Günther, S. Fritsch, neuralnet: Training of neural networks, The R journal 2 (2010) 30–38.

[35] R Core Team, R: A Language and Environment for Statistical Computing, R Foundation for Statistical Computing, Vienna, Austria, 2018. URL: https://www.R-project.org/.

[36] D. L. Doolan, C. Dobaño, J. K. Baird, Acquired immunity to malaria, Clinical microbiology reviews 22 (2009) 13–36.

[37] K. R. Dobbs, A. E. Dent, Plasmodium malaria and antimalarial antibodies in the first year of life, Parasitology 143 (2016) 129–138.

